# Association of the rs738409 polymorphism in *PNPLA3* with development and severity of non-alcoholic fatty liver disease in ethnic Bengali population of West Bengal

**DOI:** 10.1101/2024.09.02.24312965

**Authors:** Mousumi Das, Arindam Biswas, Soumik Goswami, Rajat Deb, Sukdeb Das, Debes Ray

## Abstract

**Background and Objectives:** Non-alcoholic Fatty Liver Disease (NAFLD) is a common disorder with a complex etiology. Polymorphic variant rs738409 (Ile148Met) in the *PNPLA3* gene has been reported to be associated with NAFLD among several ethnic populations. The present study aims to identify the potential association of this PNPLA3 gene variant with NAFLD among the ethnic Bengali population of West Bengal and correlate it with disease severity and biochemical parameters.

**Materials and Methods:** The Ile148Met variant was genotyped among 99 clinically diagnosed NAFLD cases and 100 ethnicity-matched controls from West Bengal using PCR-RFLP-based techniques.

**Results:** We identify statistically significant differences in genotype and allele frequencies of rs738409 between NAFLD cases and controls (P = 0.001 & 0.0057). The frequency distribution of risk genotype (GG) and risk allele (G allele) was significantly associated with a higher FIB-4 score (P = 0.0251) and Controlled Attenuation Parameter (CAP) score (P = 0.0072) but not with Liver Stiffness Measurement (LSM) value. The mean CAP score was significantly higher among the risk allele (G allele) carrier than the ‘C’ allele carrier (272.49±51.99 vs 247.38±49.46; P = 0.0328). In patients with NAFLD, the CC genotype was associated with a higher level of total bilirubin (P = 0.0189).

**Conclusion:** The Ile148Met variant of the *PNPLA3* gene is a predisposing risk factor for the development of NAFLD and is associated with increased severity of hepatic steatosis among the ethnic Bengali population of West Bengal.

## INTRODUCTION

Non-alcoholic fatty liver disease (NAFLD) is characterized by hepatic fat accumulation, with the disease spectrum ranging from simple steatosis to non-alcoholic steatohepatitis (NASH) and cirrhosis (Lazo et al., 2008). In India, the prevalence of NAFLD varies from 15% to 32% depending on the ethnic structure and study design (Bajaj et al., 2009; Mohan et al., 2009). NAFLD is a complex disease, with a substantial involvement of genetic component. Studies on twins and differences in susceptibility and progression suggest a significant heritable component to NAFLD that may be classified under the “common disease – common variant” hypothesis.

*Patatin-like phospholipase domain containing protein-3 (PNPLA3)* gene encodes a protein called adiponutrin which is found in adipocytes and hepatocytes. The function of adiponutrin is not well characterized but is thought to help regulate the production and breakdown of fats. Genome-wide association studies (GWAS) in NAFLD have identified rs738409 (c.444C>G; p.Ile448Met) in the *PNPLA3* gene to be strongly associated with hepatic fat content in the Dallas Heart Study, which included a multi-ethnic population sample (Romeo et al., 2008; Severson et al., 2016; Kozlitina et al, 2014). The rs738409 polymorphism was significantly associated with several biochemical parameters including alanine transaminase (ALT) aspartase transaminase (AST), triglycerides (TG) levels among Japanese and significantly associated with steatosis severity, hepatocellular ballooning, lobular inflammation in paediatric NAFLD cases (Oliveira et al., 2021). The rs738409 variant was also studied among Indian paticipants and data suggest that individuals harbouring the risk allele of rs738409 of *PNPLA3* are at significant risk for developing NAFLD (Sood et al., 2018; Bhatt et al., 2013). Although growing evidence suggests existence of a link between *PNPLA3* gene variants with NAFLD in different populations, data on the same and its relation to disease severity is lacking in the Bengali population of West Bengal. Therefore, in this study, we aimed to identify the potential association of the PNPLA3/ rs738409 gene variant with NAFLD among the ethnic Bengali population of West Bengal and correlate it with disease severity and biochemical parameters.

## MATERIALS AND METHODS

### Recruitment of study subjects

Patients attending the OPD of the Department of Medicine and Department of Endocrinology at the Nil Ratan Sircar Medical College and Hospital, Kolkata, West Bengal, India with ultrasound showing fatty liver were selected after exclusion of other causes. A total of 99 patients (age range 21 -73 years) were recruited in this study for the estimation of biochemical parameters (fasting plasma glucose, fasting insulin, glycated haemoglobin, serum triglyceride, HDL, LDL, total cholesterol, AST, ALT, GGT, ALT, total bilirubin, etc.), haematological parameters (platelet count), Transient Elastography (TE) and genetic analysis. The demographic details are mentioned in Table 1. The Ethics Committee of the above-mentioned Institute approved the study protocol. Informed consent was taken as per guidelines of the Indian Council of Medical Research, National Ethical Guidelines for Biomedical and Health Research involving human participants, India.

**Table 1.**
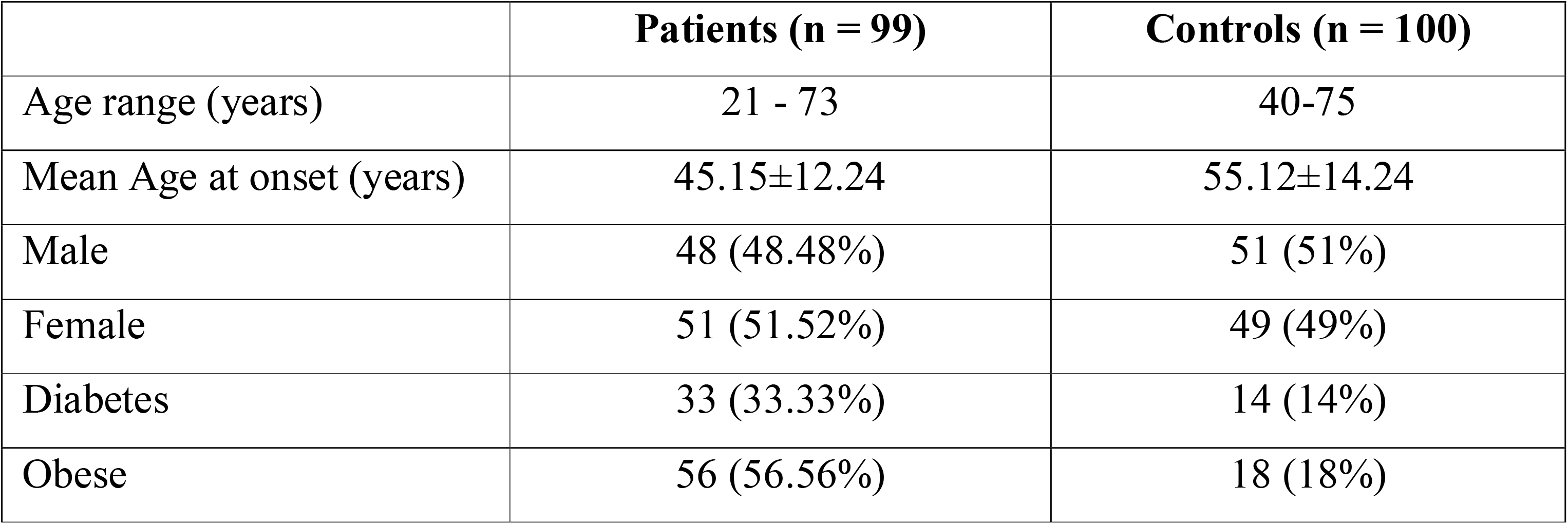
Demographic details of the studied population.

|  | **Patients (n = 99)** | **Controls (n = 100)** |
| --- | --- | --- |
| Age range (years) | 21 - 73 | 40-75 |
| Mean Age at onset (years) | 45.15±12.24 | 55.12±14.24 |
| Male | 48 (48.48%) | 51 (51%) |
| Female | 51 (51.52%) | 49 (49%) |
| Diabetes | 33 (33.33%) | 14 (14%) |
| Obese | 56 (56.56%) | 18 (18%) |

### Collection of blood samples and determination of biochemical parameters &genomic DNA isolation

Approximately, 10 mL of peripheral venous blood samples were collected and divided into two parts -one part was used for biochemical analysis and another part used for isolation of Genomic DNA using QIAamp DNA Blood Mini Kit (Qiagen GmbH, Hilden, Germany) according to the manufacturer’s instructions and dissolved in TE(10 mM Tris− HCl, 0.1 mM EDTA, pH 8.0) and stored at 4^0^C.

### Biochemical analysis

Fasting plasma glucose (FPG), glycated haemoglobin (HbA1C), serum total cholesterol, triglyceride, HDL, LDL, total bilirubin, alkaline phosphatase, AST, ALT, and GGT, levels were analyzed using Roche Cobas c-501 module Automatic Analyzer (Roche Diagnostics, Basel, Switzerland). Fasting insulin was measured using commercially available Elecsys Insulin ECLIA kits in Roche Cobas e-601 module Automatic Analyzer (Roche Diagnostics, Basel, Switzerland). Platelet count was analyzed using Sysmex Automated Hematology Analyzer XN-1000. The insulin resistance of patients was evaluated via the homeostasis model assessment of insulin resistance (HOMA-IR): HOMA-IR= fasting plasma glucose (FPG)(mg/dl) x fasting insulin (FINS)(mIU/l) /405.

### Imaging

Controlled Attenuation Parameter (CAP) and Liver Stiffness Measurement (LSM) were measured by Transient Elastography (TE). TE was performed by a single operator with Fibroscan ® Mini+ 430 model by Echosens on the right lobe of the liver, through intercostal spaces, with the participant lying in maximal abduction. According to the instructions from the manufacturer, either an M-or XL-probe was applied. CAP provides the measurement of liver ultrasonic attenuation (go and return path) at 3.5MHz on the signals acquired by Fibroscan®. It is expressed in decibels per meter (dB/m) and the final liver stiffness result was expressed in kilopascal (kPa). Both CAP and LSM were obtained simultaneously and in the same volume of parenchyma. Based on the CAP score set by the manufacturer (Fibroscan by Echosens), liver steatosis is graded as Grade 0 to Grade III-< 237dB/m: normal liver, 238-259dB/m: Grade I liver steatosis, 260-292 dB/m: Grade II liver steatosis, >292 dB/m: Grade III liver steatosis. Liver stiffness measurement (LSM) >= 8.0 kPa was considered as the cutoff level for CSLF and LSM>= 13.0 kPa for Cirrhosis. This cutoff level was chosen as it yields high positive predictive values (PPVs) for the presence of clinically significant fibrosis in previous studies (Koehler et al., 2016).

### Screening of Ile148Met (rs738409) variant in PNPLA3 gene

Peripheral blood samples were collected from study subjects and genomic DNA was isolated. The targeted region harboring the nucleotide change was amplified by PCR, digested with NlaIII (New England Biolabs, USA) as per conditions specified by the manufacturer, and electrophoresed on a 7% polyacrylamide gel.

### Statistical analysis

Biochemical parameters and comparison of clinical parameters were analyzed by Mann Whitney U test (Wilcoxon rank-sum) and Fischer exact P-value.

## RESULTS

### Genetic analysis

A total of ninety-nine patients with a mean age of 45.15±12.19 (age range 21-73 years) and one hundred ethnically matched controls free of any hepatological diseases (mean age 52.21±8.33 years) were genotyped for Ile148Met variants (rs738409) in *PNPLA3* gene. The genotype distribution of the studied variant was in Hardy-Weinberg equilibrium for the control population (P = 0.5878, chi-square = 0.2937) and deviated in the patient population ((P = 0.00045, chi-square = 12.312). The ‘G’ allele (P = 0.0057, OR = 1.839; 95% CI: 1.194 – 2.833) and ‘GG’ genotype (P = 0.001, OR = 5.429; 95% CI: 1.964 – 15.002) of rs738409 were found to be associated with the development of NAFLD among our population when compared to healthy individuals (Table 2). The risk allele (G allele) was found to be present in higher frequency among cases in comparison to controls (37.38% among cases and 24.5% among controls). Similarly, the homozygous ‘GG’, the risk genotype was also present in higher frequency among NAFLD cases (22% vs 5%), and the differences were statistically significant (Table 2).

**Table 2.** Genotype and Allele frequencies of Ile148Met (rs738409) variant in the *PNPLA3* gene and compare with clinical parameters.

|  | **CC** | **GC** | **GG** | **C** | **G** | **P-value** | **Odds ratio (95% CI)** |
| --- | --- | --- | --- | --- | --- | --- | --- |
| **Patient (n = 99)** | 47 (47.5%) | 30 (30.5%) | 22 (22%) | 124 (62.62%) | 74 (37.38%) | G vs C: **0.0057** | **1.839 (1.194 – 2.833)** |
| **Controls (n = 100)** | 56 (56%) | 39 (39%) | 5 (5%) | 151 (75.5%) | 49 (24.5%) | GG+GC vs CC: 0.2294<br>GG vs GC+CC: **0.001** | 1.408 (0.806 – 2.461)<br>**5.429 (1.964 – 15.002)** |
| **FIB-4 $\geq 1.3$ (n = 54)** | 22 (40.74%) | 16 (29.63%) | 16 (26.93%) | 60 (55.56%) | 48 (44.44%) | **0.0251** | **1.969 (1.088 – 3.564)** |
| **FIB-4 < 1.3 (n = 45)** | 25 (55.56%) | 14 (31.11%) | 6 (13.33%) | 64 (71.11%) | 26 (28.89%) | Referent |  |
| **CAP $\geq 237$ (n = 63)** | 26 (41.27%) | 18 (28.57%) | 19 (30.16%) | 70 (55.56%) | 56 (44.44%) | **0.0072** | **2.40 (1.267 – 4.546)** |
| **CAP < 237 (n = 36)** | 21 (58.33%) | 12 (33.33%) | 3 (8.34%) | 54 (75%) | 18 (25%) | Referent |  |
| **LSM > 7 (n = 42)** | 19 (45.24%) | 12 (28.57%) | 11 (26.19%) | 50 (59.5%) | 34 (40.5%) | 0.4389 | 1.258 (0.704 – 2.249) |
| **LSM $\leq 7$ (n = 57)** | 28 (49.12%) | 18 (31.58%) | 11 (19.3%) | 74 (64.91%) | 40 (35.09%) | Referent |  |
| **HOMA-IR (< 2.5), n = 24** | 12 (50%) | 5 (21%) | 7 (29%) | 29 (60.42%) | 19 (39.58%) | 0.7163 | 1.132 (0.581 – 2.205) |
| **HOMA-IR ($\geq 2.5$), n = 75** | 35 (46.67%) | 25 (33.33%) | 15 (20%) | 95 (63.33%) | 55 (36.67%) | Referent |  |
| **Obese (BMI $\geq 25$), n = 56** | 24 (42.86%) | 17 (30.34%) | 15 (26.80%) | 65 (58.04%) | 47 (41.96%) | 0.1287 | 1.580 (0.875 – 2.850) |
| **Non-Obese (BMI < 25) n = 43)** | 23 (53.49%) | 13 (30.23%) | 7 (16.28%) | 59 (68.61%) | 27 (31.39%) | Referent |  |

### Genotype to phenotype correlation

It was observed that the risk allele (G allele) was significantly overrepresented among the cases having FIB-4 score ≥ 1.3 in comparison to the cases having FIB-4 score < 1.3 (44.44% vs 28.89%; P = 0.0251; OR = 1.969;95% CI: 1.088 – 3.564) (Table 2). Similarly, the Ile148Met polymorphic variant served as a risk factor for higher CAP score (CAP score ≥ 237dB/m). The variant allele (G allele) was present among 44.44% of cases where the CAP score was ≥ 237 dB/m and 25% where the CAP score was < 237 dB/m and this difference was also statistically significant (P = 0.0072) (Table 2). Likewise, the mean CAP score was significantly higher among the risk allele (G allele) carrier than the ‘C’ allele carrier (272.49±51.99 vs 247.38±49.46; P = 0.0328) (Table 3). On the other hand, we did not find any statistically significant association between LSM with the Ile148Met variant of the *PNPLA3* gene. However, a numerically higher proportion of variant alleles was present among those cases having LSM >7 kPa when compared to LSM ≤ 7 kPa (40.5% vs 35.09%; P = 0.4389). No differences were observed for insulin resistance based on HOMA-IR value and obesity (Table 2).

**Table 3.** Comparison of clinical parameters according to PNPLA3 (rs738409; Ile148Met) variant.

| **Biochemical parameters/SNV** | **CC (n = 47)** | **GC+GG (n = 52)** | **P-value** |
| --- | --- | --- | --- |
| **FBS** | 120.37±44.87 | 112.78±44.59 | 0.1271 |
| **HBA1C** | 6.39±1.23 | 6.63±1.66 | 0.9217 |
| **Fasting insulin** | 16.96±10.47 | 15.90±8.50 | 0.7472 |
| **PLATELET(X10<sup>9</sup>/L)** | 182.44±81.65 | 179.61±79.22 | 0.9553 |
| **HOMA-IR** | 5.29±4.22 | 4.52±3.05 | 0.5791 |
| **Total Cholesterol** | 175.14±56.40 | 173.58±56.72 | 0.9079 |
| **HDL** | 46.48±13.73 | 44.57±13.42 | 0.3659 |
| **TG** | 183.43±73.72 | 157.57±70.91 | 0.0810 |
| **LDL** | 92.95±50.16 | 97.49±45.27 | 0.3868 |
| **VLDL** | 36.68±14.74 | 31.51±14.18 | 0.0810 |
| **AST(IU/L)** | 38.13±26.90 | 33.28±16.73 | 0.8457 |
| **ALT(IU/L)** | 34.19±18.74 | 32.83±16.32 | 0.9857 |
| **Total BIL(MG/DL)** | **0.76±0.51** | **0.55±0.44** | **0.0189** |
| **ALP(IU/L)** | 107.54±27.42 | 100.07±28.39 | 0.1085 |
| **GGT(IU/L)** | 32.00±25.66 | 27.89±22.85 | 0.2345 |
| **CAP** | **247.38±49.46** | **272.49±51.99** | **0.0328** |
| **LSM** | 7.33±4.97 | 7.35±4.31 | 0.8527 |
| **FIB-4** | 2.09±1.86 | 2.46±4.12 | 0.809 |

### Comparison of biochemical parameters

On comparing the biochemical parameters between the individuals having the ‘CC’ genotype (n = 47) and the individuals having ‘GC+GG’ genotypes (n = 52), a significantly higher amount of total bilirubin was found among the ‘CC’ genotype carriers than others (0.76±0.51 vs 0.55±0.44; P = 0.0189) (Table 3). Similarly, a trend of higher serum triglyceride (183.43±73.72 vs 157.57±70.91) and VLDL (36.68±14.74 vs 31.51±14.18) was observed among the ‘CC’ genotype carriers (Table 3). Other biochemical parameters like FPG, HbA1C, fasting insulin, total cholesterol, AST, ALT, ALP, and GGT were unaltered irrespective of the genotype (Table 3).

## DISCUSSION

Identification of population-specific genetic risk variants in NAFLD have long been reported in different populations including from India [Sood et al., 2018; Bhatt et al., 2013]. This variant has been consistently associated with the risk of NAFLD by genome-wide association studies as well as candidate gene studies [Zhang et al., 2015; Anstee et al., 2013; Shen et al., 2015; Singal et al., 2013; Shen et al., 2014; Zhang et al., 2014]. In the present study, we have identified p.Ile148Met (rs738409) of the *PNPLA3* gene as a risk factor for NAFLD among the ethnic Bengali population of West Bengal in both allelic and genotypic manner with odds ratio 1.839 and 5.429 respectively. The risk ‘GG’ genotype was more prevalent among the NAFLD cases compared to the healthy population (22% vs 5%), suggesting that the ‘GG’ genotype exerts a strong influence on liver fat accumulation.

The PNPLA3 is a 481 amino acids protein, also known as adiponutrin or calcium-dependent phospholipase A2 epsilon, and belongs to the PNPLA family, highly expressed in the liver. The missense variant, Ile148Met of *PNPLA3* causes a loss of function that predisposes to steatosis as a result of decreased hydrolysis of triglycerides in hepatocytes [Wilson et al., 2006; Kienesberger et al., 2009; Kollerits et al., 2009]. A mutation in the *PNPLA3* gene (adiponutrin, rs738409 C>G) encoding the substitution of isoleucine for methionine at positions 148 has been linked to the incidence and severity of alcoholic liver disease, non-alcoholic fatty liver disease, chronic hepatitis, fibrosis, and cirrhosis, as well as their complications, including hepatocellular carcinoma [Romeo et al., 2008; Valenti et al., 2010; Huang et al., 2011; Liu et al., 2014; Kalia et al., 2016; Grimaudo et al., 2020]. There are conflicting data regarding the association between the Ile148Met genotype and abnormalities in biochemical parameters including BMI, hypertriglyceridemia, and elevated AST levels in NAFLD. Kollerits et al., 2009 reported that there was a strong association of rs738409 with age, gender, ALT, lipoprotein concentration, total cholesterol, HDL, and LDL levels [Kollerits et al., 2009]. In our study, we also found a lower level of triglyceride and VLDL concentration among the individuals having the ‘GC+GG’ genotype of rs738409. Previously, Shao et al did not find any correlation between rs738409 with total bilirubin levels. In this study, we identify a decrease in total bilirubin levels among the variant allele (G allele) carriers in comparison to ‘CC’ carriers.

We found a significant association of CAP score with rs738409 of the PNPLA3 gene. The variant allele carriers had a significant increase in CAP score than the wild type. Similarly, a significantly higher frequency of risk alleles was present among the patients having higher FIB-4 scores and CAP scores. Pennisi et al., 2021 reported that the rs738409 variant is an independent risk factor for liver fibrosis progression. Patients carrying the variant allele had a significantly higher rate of fibrosis progression [Pennisi et al., 2021].

This study has several limitations including (a) a small sample size (b) samples from only one ethnic population being involved (c) the diagnosis of NAFLD was not based on liver biopsy and. Therefore, revalidation of the identified correlation with other ethnic populations is necessary.

In conclusion, the *PNPLA3* gene variant (rs738409) was associated with an increased risk of NAFLD among the ethnic Bengali population of West Bengal. The risk ‘G’ allele also positively correlated with the CAP score, thereby implying an increased risk of hepatic steatosis severity.

## Acknowledgement

The authors thank the patients who participated in the study.

## Credit authorship contribution statement

MD, AB, SG, and DR were responsible for the concept, study design, sample collection, clinical diagnosis, experimental work, data analysis and manuscript preparation. RD and SD were responsible for clinical diagnosis and manuscript preparation. All authors read the draft, provided their inputs, and agreed on the final version of the manuscript.

## Ethics statement

All procedures performed in studies involving human participants were in accordance with the ethical standards of the Nil Ratan Sircar Medical College & Hospital, Kolkata, India. The Ethics Committees (Institutional Ethics Committee, Nil Ratan Sircar Medical College & Hospital) of the abovementioned Institutes approved the study protocol. Informed consent was taken as per guidelines of the Indian Council of Medical Research, National Ethical guidelines for Biomedical and Health research involving human participants, India.

## Informed consent

Informed consent from all the participants was received before clinical data and sample collection.

## Data Availability Statement

The data described in this study are available from the corresponding author upon reasonable request.

